# Concordance of B and T cell responses to SARS-CoV-2 infection, irrespective of symptoms suggestive of COVID-19

**DOI:** 10.1101/2022.02.03.22270393

**Authors:** Marc F. Österdahl, Eleni Christakou, Deborah Hart, Ffion Harris, Yasaman Shahrabi, Emily Pollock, Muntaha Wadud, Tim D. Spector, Matthew A. Brown, Jeffrey Seow, Michael H. Malim, Claire J. Steves, Katie J. Doores, Emma L Duncan, Timothy Tree

## Abstract

**Objectives:** To assess T cell responses in individuals with and without a positive antibody response to SARS-CoV-2, in symptomatic and asymptomatic individuals during the COVID-19 pandemic.

**Methods:** Participants were drawn from the TwinsUK cohort, selected according to a) presence or absence of COVID-associated symptoms (S+, S-), logged prospectively through the COVID Symptom Study app, and b) Anti-IgG Spike and anti-IgG Nucleocapsid antibodies measured by ELISA (Ab+, Ab-), during the first wave of the UK pandemic. T cell helper and regulatory responses after stimulation with SARS-CoV-2 peptides were assessed.

**Results:** 32 participants were included in final analysis. 14 of 15 with IgG Spike antibodies had a T cell response to SARS-CoV-2-specific peptides; none of 17 participants without IgG Spike antibodies had a T cell response (Chi-squared 28.2, p<0.001). Quantitative T cell responses correlated strongly with fold-change in IgG Spike antibody titre (rho=0.79, p<0.0001) but not to symptom score (rho=0.17, p=0.35).

**Conclusions:** Humoral and cellular immune responses to SARS-CoV-2 are highly correlated, with no evidence that cellular immunity differs from antibody status four months after acute illness.

## Introduction

The COVID-19 pandemic caused by SARS-CoV-2 has been catastrophic to human health, causing over 4 million deaths worldwide by July 2021 (1). Key to controlling SARS-CoV-2 spread is the ability to identify accurately individuals with current or past infection, determining quarantine and contact tracing requirements. Current infection with SARS-CoV-2 is diagnosed using PCR (detecting viral RNA) or lateral flow antigen testing); prior infection is typically diagnosed by demonstrating a memory immune (typically antibody) response to SARS-CoV-2.

The adaptive immune response to SARS-CoV-2 comprises humoral and cell-mediated components. The humoral (or antibody) response is detectable in convalescent sera approximately 2-3 weeks after infection, with an initial IgM response followed within days by an IgG response, including against the Spike protein (also the target of vaccines (2)) and, less specifically, against the Nucleocapsid protein. Additionally, a neutralising antibody response may be measurable, which assesses the functional capacity of a convalescent serum to inhibit virus infection *in vitro* (3). Complementary to the antibody response is a cellular response driven by T cells, particularly CD4^+^ T helper cells. When stimulated by a pathogen, naïve CD4^+^ cells differentiate into T helper cell subsets which orchestrate the immune response, including supporting pathogen-specific cytotoxic (CD8^+^) T cells, and stimulating B-cells to produce a high affinity pathogen-specific antibody response. Other CD4^+^ T cells differentiate into T regulatory cells which attenuate immune responses to the pathogen(4). Following clearance of infection, pathogen-specific memory T cell responses play an important role in protective immunity, and are detectable in peripheral blood long-term.

Antibody responses have been detected in most individuals after acute COVID-19 (3) (5). However, some individuals reporting symptoms suggestive of COVID-19, including individuals reporting prolonged symptom duration suggestive of the Post-COVID Syndrome (“Long COVID “), do not have detectable antibody responses (6). One possible explanation is infection with other respiratory pathogens (such as influenza virus) whose symptom profile overlaps with COVID-19. This was particularly evident in the first wave of the pandemic; however, as the pandemic progressed, with introduction of social distancing and personal protection, circulation of other respiratory pathogens declined and SARS-CoV-2 became the dominant respiratory infection (7). Another possibility is false negative testing, as thresholds for defining an antibody response as positive or negative reflect a compromise between assay sensitivity and specificity, neither being 100%. In addition, antibody responses to SARS-CoV-2 decline over time, also observed with other coronaviruses including SARS-CoV-1 (8). In contrast, T cell responses are usually prolonged and, although their frequency may wane with time, can be demonstrable years after initial infection - for example, T cell responses following SARS-CoV-1 infection are detectable for over 17 years (to date) [(9)].

Here we assess humoral and cell-mediated responses to SARS-CoV-2, in symptomatic and asymptomatic individuals during the first UK wave of the COVID-19 pandemic. In particular, we assess whether individuals symptomatic of COVID-19 but without a detectable antibody response have demonstrable cell-mediated immunity.

## Materials and Methods

### TwinsUK

The TwinsUK cohort is the largest community-based cohort of adult twins in the UK, with >14 000 registered individuals (>7 000 pairs) assessed longitudinally over nearly 30 years. Their experience of the COVID-19 pandemic was closely monitored, with 10 230 individuals participating in regular questionnaires about prior symptoms (10),(11). Of these, 431 individuals participated in a home visit study during May-June 2020 (study protocol, participant demographics, and inclusion/exclusion criteria previously published (6)). Briefly, participants were selected based on: (a) proximity of both twins (within 80 miles of St Thomas ‘ Hospital, Westminster); (b) sufficient symptom reporting in the COVID Symptom Study (discussed below) to enable calculation of a COVID ‘symptom score ‘ (12); and (c) availability during the study period. Serum samples were tested for IgG antibody against SARS-CoV-2 Spike protein.

Subsequently, individuals were selected to form four groups, defined by symptom score and IgG Spike antibody responses from the initial home visit: symptom-positive, antibody-positive; symptom-positive, antibody-negative; symptom-negative, antibody-positive (i.e., asymptomatic infection); and symptom-negative, antibody-negative (i.e., control group). Participants were then revisited, to collect PBMCs and a contemporaneous serum sample for repeat antibody testing.

### The COVID Symptom Study (CSS)

The CSS was launched jointly on 24 March 2020 by ZOE Limited and academics of King ‘s College London, Massachusetts General Hospital, and Lund and Uppsala Universities, through a smart phone application(12). Briefly, on registration participants provide baseline demographic data and subsequently are prompted daily to report their health status (including being asymptomatic), health care access, vaccination, testing, etc. Data on key symptoms is combined into a ‘symptom score ‘ from 0 to 1.0, with a score above 0.5 defining probable COVID-19 (here, “Symptom-Positive “). During the first UK wave, this model showed 65% sensitivity and 78% specificity for self-reported SARS-CoV-2 infection (defined by reverse transcription PCR) [(12)]. Many of the TwinsUK cohort also participate in the CSS, with linkage of their data.

### Humoral assays

Spike and Nucleocapsid protein were expressed as previously described (13). All sera were heat-inactivated at 56°C for 30 mins before use. High-binding ELISA plates (Corning, 3690) were coated with antigen (Spike or N) at 3 µg/mL (25 µL per well) in PBS, either overnight at 4°C or 2 hr at 37°C. Wells were washed with PBS-T (PBS with 0.05% Tween-20) and then blocked with 100 µL 5% milk in PBS-T for 1 hr at room temperature. Wells were emptied and sera (diluted at 1:50 in milk) added and incubated for 2 hr at room temperature. Control reagents included CR3009 (2 µg/mL), CR3022 (0.2 µg/mL), negative control plasma (1:25 dilution), positive control plasma (1:50) and blank wells. Wells were washed with PBS-T. Secondary antibody was added and incubated for 1 hr at room temperature. IgM was detected using Goat-anti-human-IgM-HRP (1:1,000) (Sigma: A6907) and IgG was detected using Goat-anti-human-Fc-AP (1:1,000) (Jackson: 109-055-098-JIR). Wells were washed with PBS-T and either AP substrate (Sigma) was added and read at 405 nm (AP) or 1-step TMB substrate (Thermo Scientific) was added and quenched with 0.5 M H2S04 before reading at 450 nm (HRP). For binary classification, we used a 4-fold increase above background in both IgG Spike (S) and Nucleocapsid (N) antibody titre to define a case as “Antibody-Positive “, based on previously established thresholds (13). For continuous variable analyses, the fold-change in IgG Spike titre was used.

### Analysis of T Cell Responses

PBMCs were isolated from Li Hep blood by density gradient centrifugation using Lymphoprep (Axis Shield), cryopreserved in CS10 n CryoStor® (Sigma-Aldrich) and stored in in vapour phase liquid nitrogen. Cryopreserved PBMCs were thawed, and viability assessed by trypan blue exclusion following a resting period.

### PBMC stimulation with SARS-CoV-2 overlapping peptide pools

PBMC were stimulated with pools of overlapping peptides spanning the whole sequence of the SARS-CoV-2 Matrix (M) and Nucleocapsid (N) proteins (Peptivator peptide pools, Miltenyi) and two pools spanning the S1 and S2 domains of the SARS-CoV-2 spike protein (Peptivator_Prot_S1 Miltenyi and PepMix SARS-CoV-2 vial 2, JPT Peptide Technologies). These peptide pools can stimulate both MHC-I and MHC-II restricted T cells without HLA bias (14). Response to the S1 protein subunit is most comparable with IgG-S antibody testing. All peptides were used at a final concentration of 0.33 ng/µl. Superantigen Enterotoxin B (SEB) at 100ng/mL (Sigma Aldrich) was used as a positive control; Infanrix, a hexa-vaccine (GlaxoSmithKline) and Influvac, an Influenza surface antigen vaccine (Abbott Biologicals) were combined (HA + INF) and used to examine anamnestic responses induced by vaccination or infection. Peptide diluent (DMSO) was used as a negative control. PBMC (1-2×106 /stimuli) were incubated for 18h at 37°C in 48-well plates in X-Vivo media (Lonza) supplemented with 5% human AB serum (Sigma) and 0.4μg/mL anti-CD40 antibody (BioXcell).

### Flow Cytometry

Following incubation, PBMC were stained with a live/dead cell marker (LIVE/DEAD™ Fixable Near-IR Dead Cell Stain Kit - Invitrogen) and cell surface markers (Supplementary Table 1). Samples were acquired on a LSRFortessa Flow Analyser (BD Biosciences) and analysed using the software FlowJo (TreeStar Inc., version 10.7.2). As previously described, we defined activated conventional helper T cells based on the expression of CD69 and CD40L (CD154) and activated regulatory T cells based on upregulation of 4-1BB (CD137) and GARP in CD40L negative cells (15–17).

**Table 1:** Demographic data of participants, by grouping.

|  | **Total Cohort** | **Ab+ S+** | **Ab+ S-** | **Ab- S+** | **Ab-S-** |
| --- | --- | --- | --- | --- | --- |
| **Total n** | 32 | 9 | 6 | 12 | 5 |
| **Age - Mean (SD)** | 44.2 (13.4) | 44.6 (9.0) | 46.2 (19.5) | 40.8 (12.3) | 49.2 (16.2) |
| **Sex (% female)** | 87% | 67% | 100% | 92% | 100% |

Abbreviations: Ab+ antibody-positive; Ab- antibody-negative; S+, symptom-positive; S-, symptom-negative. SD=standard deviation

Antigen-specific T cell responses were described as the frequency of cells responding to each stimulus as a percentage of live total CD4^+^ T cells following subtraction of unstimulated controls. Negative values were set to zero.

Stored PBMCs from anonymised healthy controls recruited pre-pandemic were used to define thresholds for T cell responses to SARS-CoV-2 peptides in ROC analysis. A threshold of 0.22% increase in frequency of live T cells responding to SARS-CoV-2 peptide pools was established as optimal (sensitivity 76.9%, specificity 80%) (Figure 1A).

**Figure 1:**
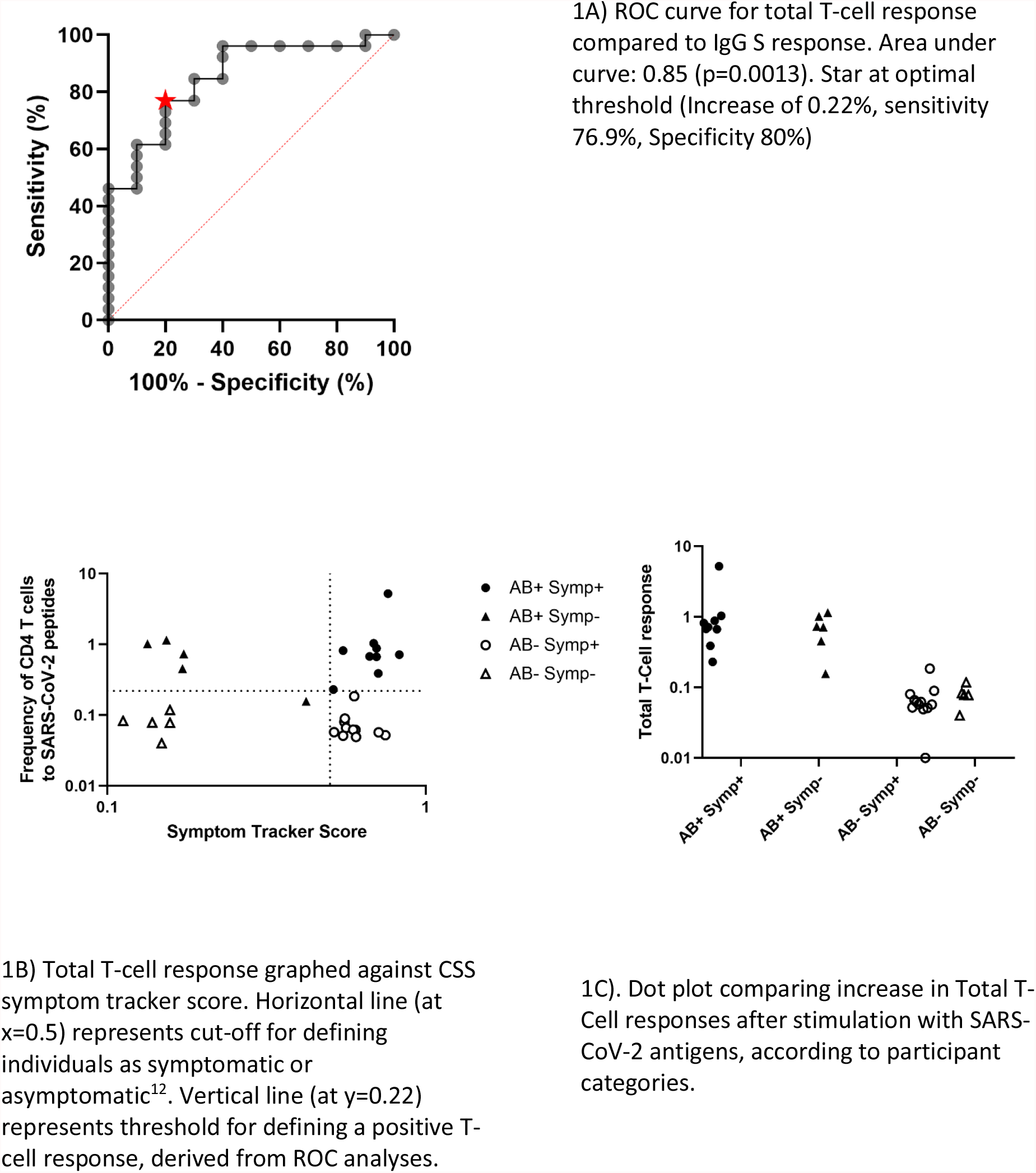
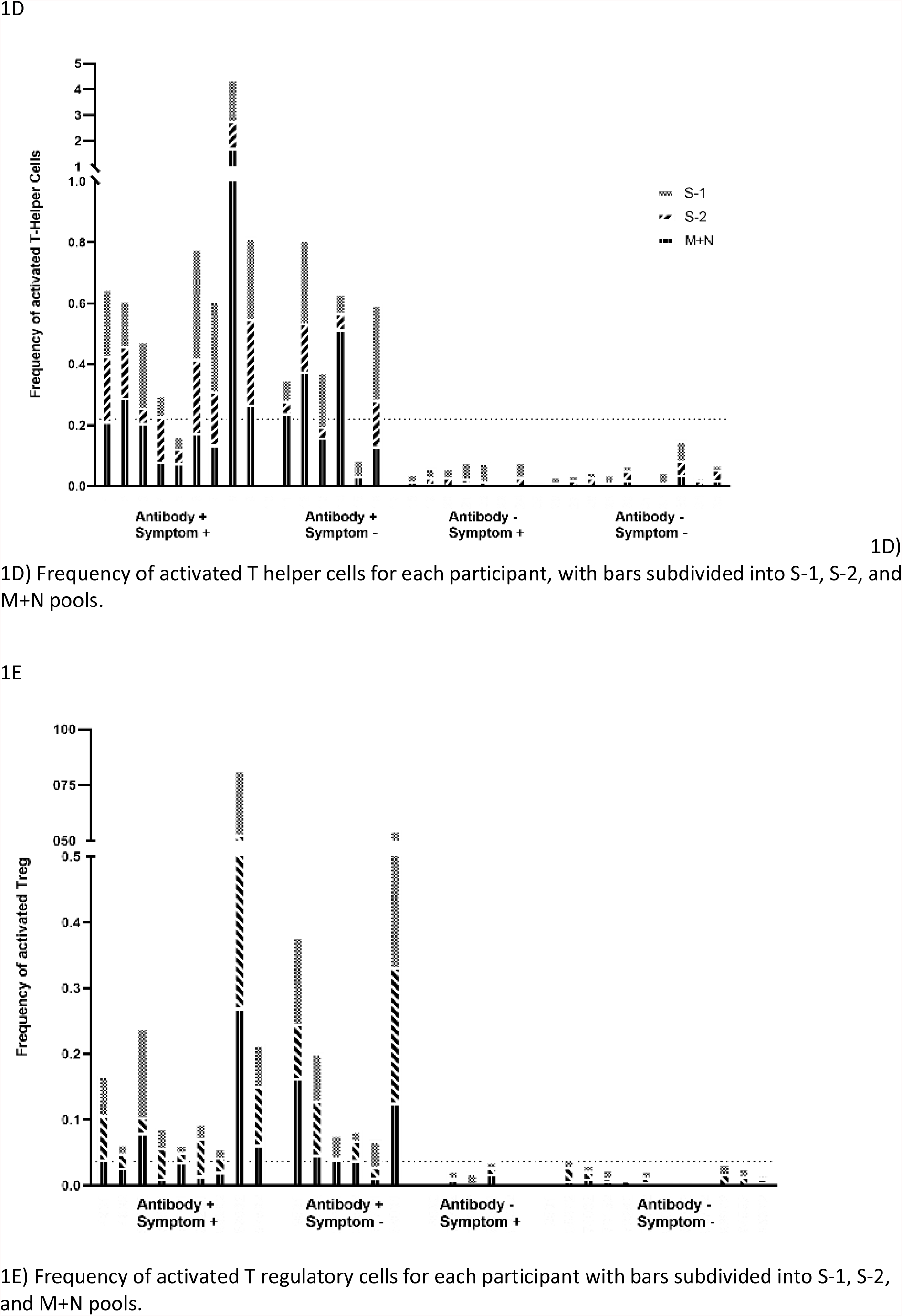
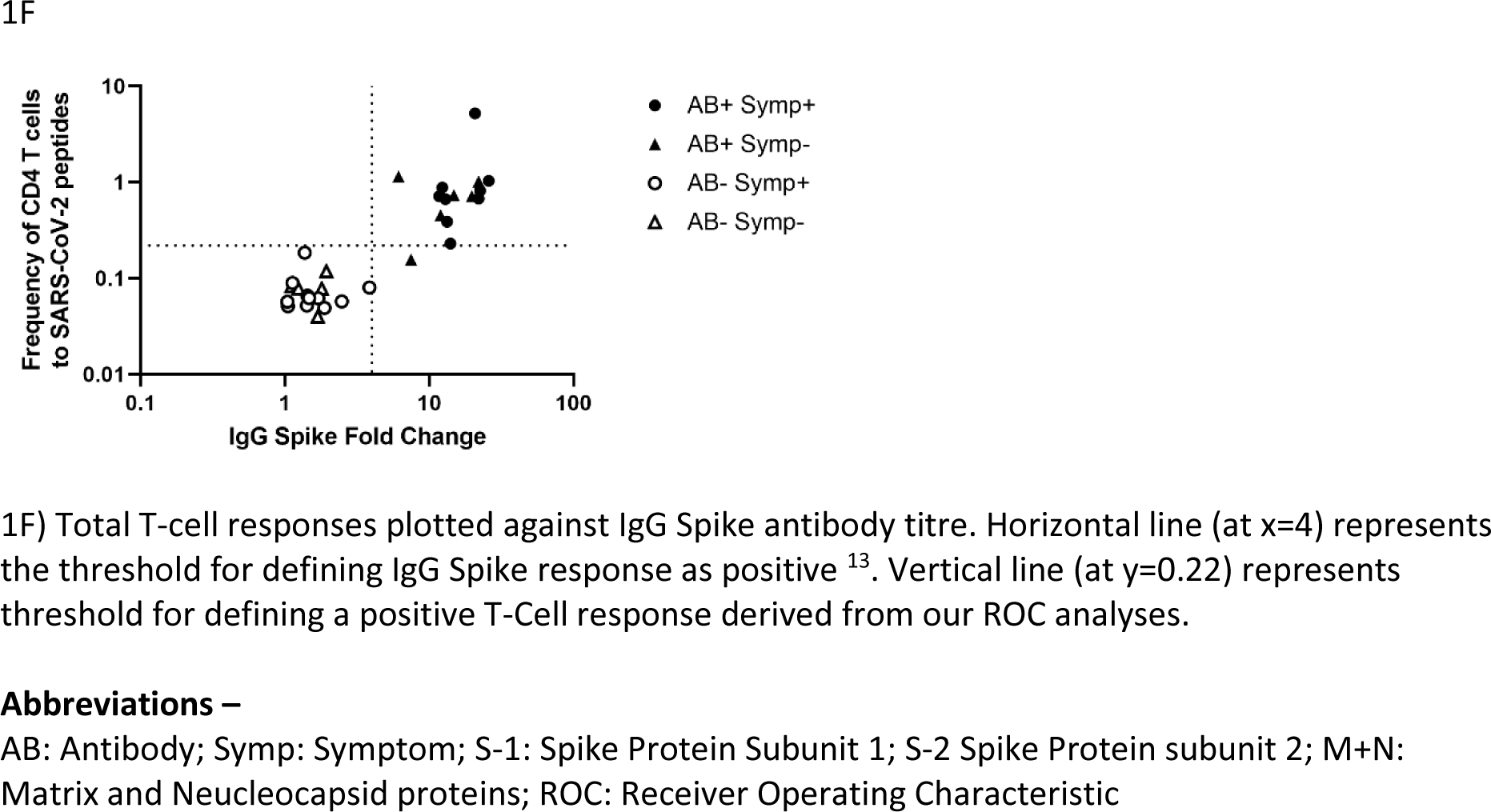
T-cell responses according to antibody and symptom status.

### Statistical Methods

Participant ascertainment is descriptive. The likelihood ratio from receiver operating characteristic (ROC) curve analysis was used to define the optimal threshold to differentiate positive vs. negative total T cell responses, as well as T helper and T regulatory responses individually. Associations between binary thresholds of IgG Spike antibody status and combined T cell responses were assessed with Chi-squared testing. When analysed as continuous variables, antibody response, symptom score and T cell responses were assessed using Wilcoxon-Rank sum testing. Spearman Rank correlation coefficients were calculated for overall and subclass T cell responses after stimulation with antibody responses, and with symptom score (both as continuous variables).

### Ethics

The TwinsUK study was approved by NHS London – London-Westminster Research Ethics Committee (REC reference EC04/015), and Guy ‘s and St Thomas ‘ NHS Foundation Trust Research and Development (R&D). The TwinsUK Biobank was approved by NHS North West - Liverpool East Research Ethics Committee (Reference: 19/NW/0187), IRAS ID 258513. All participants provide written, informed consent. The CSS was approved by KCL Ethics Committee (REMAS ID 18210, review reference LRS-19/20-18210). Upon registration, all subscribers provide consent for their data to be available for COVID-19 research. Participant samples for the ROC analysis were originally approved for use by the London-Bromley Research Ethics Committee (Reference: 08/H0805/14), with extension permitted under the extended Control of Patient Information (COPI) notice 2020/21 and specific approval from the Committee Chair for this study.

## Results

### Description of Cohort

Of 431 individuals taking part in the home visit study, 384 had also participated sufficiently in the CSS to allow calculation of a symptom score (6). Participation in the current study is outlined in Supplementary Figure 1. Thirty-four individuals had a symptom score predictive of COVID-19 (‘symptom-positive ‘). Twelve of 34 could not be revisited (moved out of defined range, declined further involvement, did not respond to contact, or were symptomatic for COVID-19 at time of planned repeat visit, precluding research team attendance). Of the remaining 22 in this group, two of the 15 who were IgG Spike antibody-negative at first home visit had seroconverted on re-assessment. A further two symptomatic but initially antibody-negative participants were excluded as repeat testing yielded inconsistent antibody results.

Twelve asymptomatic participants were positive for both IgG Spike and Nucleocapsid antibodies. Six could not be re-visited for one of the above reasons. Six asymptomatic antibody-negative individuals were chosen as controls, based on proximity to other study participants (to minimise travel by the research team); however, one proved symptom-positive on visiting and was reclassified.

Thus, final numbers in the symptomatic group were 12 antibody-negative and 9 antibody-positive individuals, and in the asymptomatic group 5 antibody-negative and 6 antibody-positive individuals. Demographic information on these participants within these final groupings is shown in Table 1. In symptomatic individuals, median time from symptom onset to PBMC collection and repeat serology was 123 days (IQR 111-130 days).

### T cell Responses vs. Symptom Scores

There was no association between T cell responses to SARS-CoV-2 peptides and either binary symptom status (Table 2; Chi-squared 0.40 p=0.529) nor correlation with actual symptom score (p>0.05 for all analyses; Table 3 Figure 1F). This was true for both T-Helper and T-Regulatory cells, as well as additional cell surface markers tested.

**Table 2:** Relationship between T cell responses, IgG Spike antibody status and symptom status.

| **IgG Spike antibody status** | **Combined T Helper + T Regulatory Responses** |  |  |
| --- | --- | --- | --- |
|  | Positive | Negative | Total |
| **Positive** | 14 | 1 | 15 |
| **Negative** | 0 | 17 | 17 |
| **Total** | 14 | 18 | 32 |
| **Symptom Status** |  |  |  |
| **Positive** | 9 | 12 | 21 |
| **Negative** | 5 | 6 | 11 |
| **Total** | 14 | 18 | 32 |

**Table 3:** Correlation of quantitative T cell responses (overall and by subtype) after stimulation with SARS-CoV-2 peptides, with IgG Spike antibody fold change, and CSS app symptom score.

|  | Correlation with Ab Response |  | Correlation to Symptoms |  |
| --- | --- | --- | --- | --- |
|  | Spearman's Rho | p-value | Spearman's Rho | p-value |
| **T Helper response:** |  |  |  |  |
| All SARS-CoV-2 peptides | 0.83 | $p < 0.0001$ | 0.16 | $p = 0.39$ |
| S1 Subunit | 0.74 | $p < 0.0001$ | -0.04 | $p = 0.83$ |
| S2 subunit | 0.71 | $p < 0.0001$ | 0.27 | $p = 0.14$ |
| M+N subunits | 0.83 | $p < 0.0001$ | 0.10 | $p = 0.6023$ |
| HA + INF | 0.02 | $p = 0.91$ | 0.55 | $p = 0.0018$ |
| SEB | 0.17 | $P = 0.37$ | 0.39 | $p = 0.0315$ |
| **T Regulatory response:** |  |  |  |  |
| All SARS-CoV-2 peptides | 0.63 | $p = 0.0001$ | 0.22 | $p = 0.23$ |
| S1 Subunit | 0.48 | $P = 0.006$ | 0.02 | $p = 0.92$ |
| S2 subunit | 0.49 | $p = 0.005$ | 0.31 | $p = 0.95$ |
| M+N subunits | 0.68 | $p < 0.0001$ | -0.06 | $p = 0.76$ |
| HA + INF | -0.11 | $p = 0.54$ | 0.46 | $p = 0.0096$ |
| SEB | 0.05 | $p = 0.79$ | 0.26 | $p = 0.16$ |
| **Combined T cell Response:** |  |  |  |  |
| All SARS-CoV-2 peptides | 0.79 | $p < 0.0001$ | 0.17 | $p = 0.35$ |

Abbreviations - S1: Spike Protein subunit 1; S2: Spike Protein subunit 2; M+N Subunits: Matrix and Nucleocapsid protein complex; HA + INF: Infrarix Hexa-vaccine and Influvac antigens; SEB: Superantigen Enterotoxin B

### T cell responses and antibody status

None of the 17 antibody-negative participants showed a T cell response to antigen pools spanning Matrix, Nucleocapsid, and the S1 and S2 domains of Spike. In contrast, 14 of 15 antibody-positive individuals demonstrated a clear T cell response (Chi-squared=28.2, p<0.001) (categorical data shown in Table 2; qualitative T cell responses shown in Figures 1B, 1C, 1D).

### Correlations between T cell responses and IgG Spike Antibody Titre

IgG Spike antibody titre correlated strongly with T cell responses to all SARS-CoV-2 antigen pools, considered as T cell responses overall (rho= 0.79, p<0.0001; Figure 1E) and as T helper and T regulatory responses individually (correlation of IgG Spike antibody titre with T helper responses: rho= 0.83, p<0.0001; and with T regulatory responses: rho=0.63, p=0.0001, Table 3).

### Associations with Control Antigens

There was no correlation between IgG Spike antibody level and T cell responses (overall or by subtype) to control antigens (HA+INF and SEB). Symptom score correlated with both T helper and T regulatory responses to HA + INF antigen stimulation (Table 3), however this was only in those who were also IgG Spike antibody positive (rho=0.81, p=0.0005) and not seen in those without IgG Spike antibodies (rho=0.33, p=0.20).

## Discussion

Here we have shown strong association and correlation between IgG Spike antibody responses and T cell responses to SARS-CoV-2 peptides. Correlation of IgG-S antibody titres were higher for T helper responses compared with T regulatory cells, which is unsurprising given the role of T helper cells in generation of B cell antibody responses. However, strong correlation was observed between T helper and T regulatory responses. We saw no evidence that individuals without a humoral response had a cellular response to SARS-CoV-2 infection, irrespective of symptoms suggestive of COVID-19.

Currently, the main use of IgG Spike antibody testing is to assess for previous infection and/or vaccine immunogenicity. Our data suggest that testing of T cell responses is unlikely to add to the information gained from antibody testing, at least in the first few months after infection. Antibodies against SARS-CoV-2 antigens decline over time (14); the close relationship we have observed between IgG Spike antibody titre and T cell reactivity may differ at later time points post infection, noting that T cell responses after SARS-CoV-1 infection can be extremely long-lasting (9). The relationship between antibody responses after vaccination (which also decline with time (18)) and protection against SARS-CoV-2 infection is an active topic of research. Whether maintaining some threshold antibody value is necessary or whether vaccination-induced cellular responses are sufficient for immunity is not currently known. However, our data could inform future research on the post-COVID syndrome (19), as T cell responses could be used to confirm previous infection for individuals with long symptom duration who lack other evidence of prior infection with SARS-CoV-2(20) (i.e., detection of virus by PCR or lateral flow antigen testing, or antibody responses), although our methodology would not readily transfer to a clinical setting and the effect of vaccination would again need consideration in interpreting results.

Our data also emphasise that the symptoms of SARS-CoV-2 can overlap with other illnesses including infection with other respiratory viruses, given the correlation between symptoms and T cell responses to HA + INF stimulation, in several individuals who were also positive for IgG spike antibodies. Whilst this could indicate previous vaccination against influenza virus, it may also indicate that some symptomatic individuals had influenza as well as SARS-CoV-2 infection; and indeed whether these individuals ‘ symptoms were due only to SARS-CoV-2 *per se* is a moot – and untestable – hypothesis. Here we would note that 1-4% of respiratory swabs reported to Public Health England showed influenza in March and April 2020 (7) although influenza subsequently declined with social distancing measures. Symptom overlap and correct disease attribution will become more relevant as social distancing restrictions are lifted, with the expected resurgence in circulation of common viral illnesses towards more usual population prevalence (21).

In defining symptomatic groups, we used a validated algorithm for predicting COVID-19 (12), although the data from this small cohort raise some questions about the robustness of this algorithm. Although IgG Spike antibody levels decline over time after natural infection, individuals were assessed on two occasions; categorisation in this study was based on contemporaneous collection of PBMCs and serology. Our laboratory methods included externally validated and published antibody testing methodology (13); and we parsed T helper and T regulatory cell responses using previously published methodologies (15, 22). However, our sample size is small (in part due to travel restrictions) and predominantly female (reflecting the nature of the TwinsUK cohort). Here we note that males are more severely affected by acute COVID-19 although there is no *a priori* reason to suspect concordance between IgG-Spike antibody status and T cell response would differ by gender. We did not screen our cohort for other infections such as influenza. We did not assess for an isolated CD8^+^ response; however, it has been demonstrated elsewhere that CD8^+^responses and CD4^+^ responses to SARS-CoV-2 are strongly associated(14). Whilst it would also be interesting to assess longitudinal patterns of IgG Spike and T-Cell responses to SARS-CoV-2 infection, our cohort are now vaccinated, and the overlap between responses to natural infection and vaccination precludes further analysis. Lastly, at the start of our home visit study (6) community RT-PCR testing was not routinely available in the UK: thus, we would be unable to detect infected individuals (i.e., RT-PCR-positive for SARS-CoV-2) who failed to mount either a B and/or T cell response.

## Conclusion

We have demonstrated strong correlation between IgG Spike antibody response to SARS-CoV-2 infection and T cell reactivity (both T helper and T regulatory cells) against SARS-CoV-2-derived peptides. Our study suggests that IgG Spike antibodies are sufficient indication of recent infection; but as antibody titres decline over time future research may be warranted to investigate the value of T cell responses in confirming historic SARS-CoV-2 infection. This may be of particular relevance in defining and managing individuals with the Post COVID syndrome(19), at least in a research setting.

## Data Availability

Data collected in the COVID Symptom Study smartphone application are shared with other health researchers through the UK National Health Service-funded Health Data Research UK (HDRUK) and Secure Anonymised Information Linkage consortium, housed in the UK Secure Research Platform (Swansea, UK). Anonymised data are available to be shared with researchers according to their protocols in the public interest (https://web.www.healthdatagateway.org/dataset/fddcb382-3051-4394-8436-b92295f14259).
The TwinsUK Resource Executive Committee (TREC) oversees management, data sharing and collaborations involving the TwinsUK registry (for further details see https://twinsuk.ac.uk/resources-for-researchers/access-our-data/). Data relevant to T-Cell and B-Cell testing may be discussed with the authors. 

## Abbreviations

COVID-19: Coronavirus Disease 2019
CSS: COVID Symptom Study
DMSO: Dimethyl Sulfoxide
ELISA: enzyme-linked immunosorbent assays
SARS-CoV-2: Severe Acute Respiratory Syndrome Coronavirus 2
PBMCs: peripheral blood mononuclear cells
REMAS: Research Ethics Management Application System
ROC: receiver operator characteristic

## Acknowledgements

We thank the volunteers of TwinsUK without whom this work would not be possible, and all participants who entered data into the C-19 Covid Symptom Study App. We thank the staff of the Department of Twin Research at King ‘s College London for their work in contributing to the running of the study and data collection - in particular, Samuel Wadge, Gulsah Akdag, Julia Brown, Alyce Sheedy and Rachel Horsfall for their frontline work conducing home visits and sample processing during a very challenging time; and Zoe Limited for access to the CSS app. We would also like to thank the healthy volunteers who work with King ‘s College London, for their contribution providing control sera for our ongoing studies.

