## Supplementary Table 1 + Supplementary Figure 1 for "Concordance of B and T cell responses to SARS-CoV-2 infection, irrespective of symptoms suggestive of COVID-19"

**Supplementary Table 1:** Cell Surface markers used to identify T cell subsets

| Cell Surface Marker | Clone/Fluorochrome | Manufacturer |
|---|---|---|
| **GARP** | Clone 7B11- APC | BioLegend |
| **GARP** | Clone G14D9-eFluor 660 | eBioscience |
| **CD19** | Clone HIB19 -APC/Cy7 | BioLegend |
| **CD137** | Clone 4B4-1 - BV421 | BioLegend |
| **CD134/OX-40** | Clone Ber-ACT35 – BV605 | BioLegend |
| **CD154** | Clone 24-31 - BV771 | BioLegend |
| **CD69** | Clone FN50 - FITC | BioLegend |
| **CD14** | Clone HCD14 - APC/Cy7 | BioLegend |
| **HLA-DR** | Clone L243 - PE-Dazzle 594 | BioLegend |
| **CD4** | Clone RPA-T4 - BUV395 | BD |
| **CD8** | Clone SK1 - BUV737 | BD |
| **CD45RA** | Clone HI100 - BV785 | BioLegend |
| **CCR7** | Clone 3D12 - APC-R700 | BD |
| **CCR6** | Clone 11A9 - BV650 | BD |
| **CXCR6** | Clone K041E5 - PE | BioLegend |
| **CXCR3** | Clone G025H7 - PE/Cy5 | BD |
| **CXCR5** | Clone J252D4 - PE/Cy7 | BioLegend |

**Supplementary Figure 1:** Flowchart of enrolment into cohort

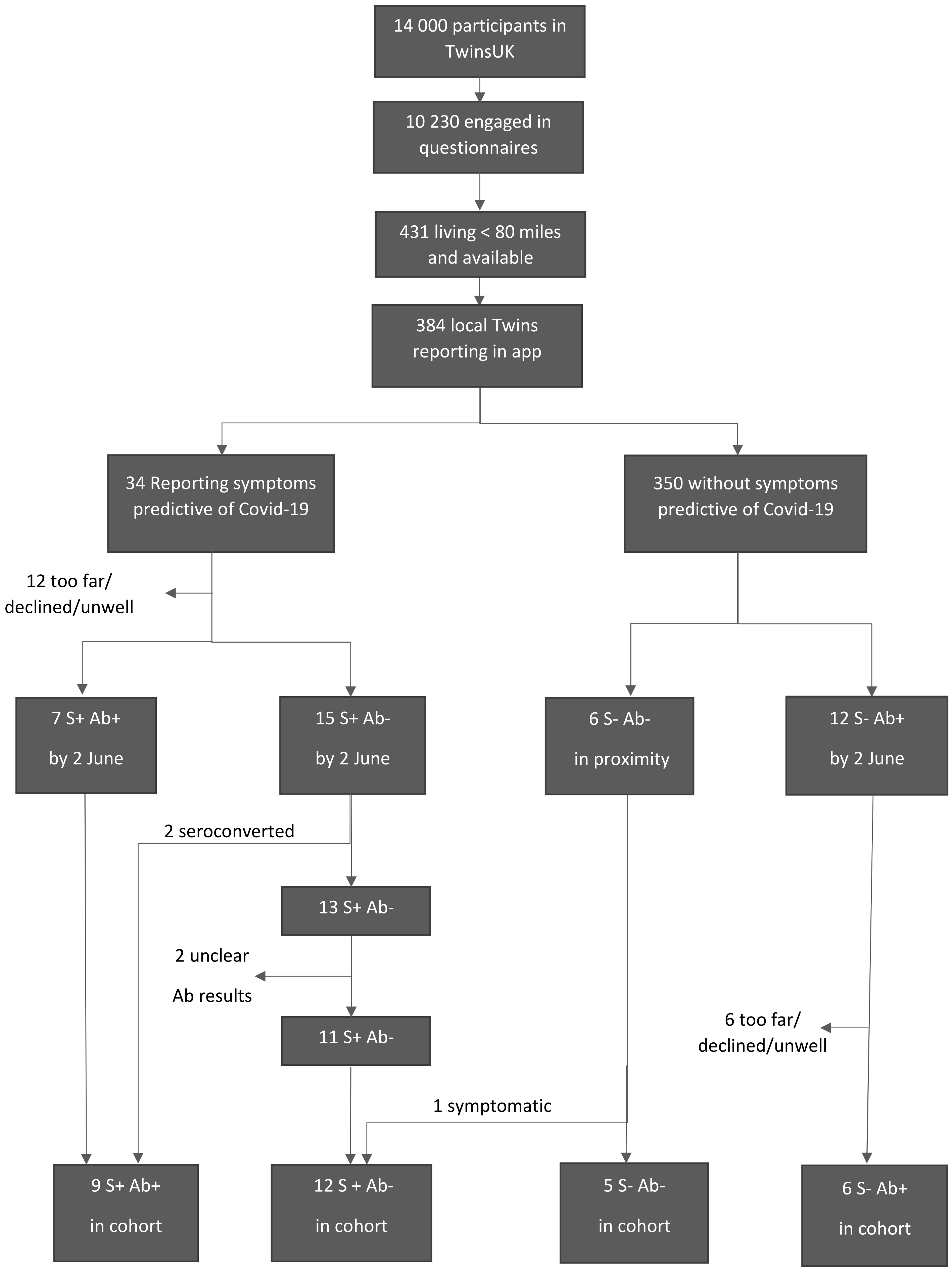
